# Viroselect: A novel SARS-CoV-2 detection assay to resolve inconclusive samples

**DOI:** 10.1101/2021.02.11.21251605

**Authors:** Ketki Jawade, Akhauri Yash Sinha, Sharad Bhagat, Shilpa Bhowmick, Bhagyashree Chauhan, Snehal Kaginkar, Harsha Palav, Nandini Kasarpalkar, Pratik Devadiga, Kalyani Karandikar, Sachee Agarwal, Jayanthi Shastri, Kiran Munne, Vikrant M Bhor, Smita D Mahale, Subhanjan Bhowmik, Dhanashree Jagtap, Vainav Patel

## Abstract

**Background:** India bears the second largest burden of SARS-CoV-2 infection. A multitude of RT-PCR detection assays with disparate gene targets including automated high throughput platforms are available. Varying concordance and interpretation of diagnostic results in this setting can result in significant reporting delays leading to suboptimal disease management. Here, we report the development of a novel ORF-1a based SARS-CoV-2 RT-PCR assay, Viroselect, showing high concordance with conventional assays and the ability to resolve inconclusive results generated during the peak of the epidemic in Mumbai, India.

**Methods:** We identified a unique target region within SARS-CoV-2 ORF1a, non-structural protein (nsp3), that was used to design and develop our assay. This hypervariable region (1933-3956) between SARS-CoV-2, SARS-CoV, and MERS-COV was utilized to design our primers and probe for RT-PCR assay. We further evaluated concordance of our assay with commonly used EUA (USFDA) manual kits as well as an automated high throughput testing platform. Further, a retrospective analysis using Viroselect on samples reported as ‘inconclusive’ during April-October 2020 was carried out.

**Results:** A total of 701 samples were tested. Concordance analysis of 477 samples demonstrated high overall agreement of Viroselect assay with both manual (87.6%; 95% CI) as well as automated (84.7%; 95% CI) testing assays. Also, in the retrospective analysis of 224 additional samples reported as ‘inconclusive’, Viroselect was able to resolve 100% (19/19) and 93.7% (192/205) samples which were termed inconclusive by manual and automated high throughput platform respectively.

**Conclusion:** We show that Viroselect had high concordance with conventional assays, both manual and automated, as well as highlight its potential in resolving inconclusive samples.

## INTRODUCTION

The striking difference between previous SARS-CoV/MERS-CoV outbreaks and the ongoing SARS-CoV-2 pandemic is the increased contagious nature of the latter (Yang et al., 2020). This has resulted in over 100 million SARS-CoV-2 infections globally and ∼2.2 million deaths to date (WHO COVID-19 Dashboard, 2021). India bears the second largest burden of infected individuals with 10 million infections and over 150,000 deaths so far (ICMR COVID-19 Testing Status, 2021). Efficient and rapid diagnosis is critical to optimal management of disease spread as well as clinical management of COVID-19. To ensure maximum coverage (WHO Feature Story, 2020) and accessibility to testing at economically feasible rates, Government of India, has successfully fostered and promoted development of indigenous diagnostic kits as well integrated cutting-edge high throughput testing facilities (Gupta et al., 2020; Poljak et al., 2020). The availability of a range of diagnostic kits with disparate viral targets has also posed a challenge in terms of concordance of results as well as inconclusive results (Bhattacharya et al., 2020; Hur et al., 2020; Pujadas et al., 2020). Also, implementation of high-throughput platform-based testing while rapidly increasing diagnostic capability, also has the potential of generating inconclusive results. As a designated national centre of excellence for COVID-19 diagnostic kit validation as well as a high throughput testing centre and having encountered the aforementioned issues, we developed an in-house RT-PCR assay, Viroselect, which was evaluated for concordance with commonly used EUA (USFDA) manual kits as well as with our high throughput COBAS 6800 testing platform. Further we assessed its utility in resolving inconclusive tests by both testing modalities.

## MATERIALS AND METHODS

### 1. Viroselect Assay Development

#### 1.1 Primer and Probe Design

A novel strategy (described in results) was used to design primers for detection of SARS-CoV-2. These sequence regions were unique to SARS-CoV-2 but had maximum variation with SARS-CoV and MERS-CoV. The primer and probe were optimized for an in-house Viroselect assay described below. All the primer and probes used in the study were synthesized from Macrogen Inc., Korea.

### 2. SARS-CoV-2 Detection

#### 2.1 RNA Extraction

Oropharyngeal/ nasopharyngeal swab samples collected from suspected COVID-19 patients in Viral Transport Medium (VTM) were used for SARS-CoV-2 detection. Viral RNA Extraction from these VTM samples, were carried out using Maverick Magnetic Bead Based Nucleic Acid Extraction Kit (Cat. No.: EC00001B, MyLabs Discovery Solutions Pvt. Ltd., Lonavla, Maharashtra, India) on Thermo Scientific KingFisher Flex Purification System and MagRNA-II Viral RNA Extraction Kit (Cat. No.: G2M030420, Genes2Me Pvt. Ltd, Gurugram, Haryana, India) on MGISP-100 automated nucleic acid extraction and purification system (MGI Tech Co., Ltd, Yantian, Shenzhen, China) as per the manufacturer’s instructions.

#### 2.2 Viroselect Assay

Biplex RT-PCR assay was conducted for Viroselect assay. ORF1a (nsp3) gene for the detection of SARS-CoV-2 Virus along with human RNase P, used as internal control, from human patient samples. The primer and probe concentrations were the following, 0.5 µmole/L for primers and 0.5 µmole/L of probe concentration for ORF1a (nsp3) and 0.5 µmole/L for primers and 0.2 µmole/L of probe concentration for Human RNaseP. The TaqMan probe was labelled with 6-carboxyfluroscien (6-Fam) 5’ and BHQ1 3’ for ORF1a and with Cy5 at 5’ end and BHQ3 at 3’ end for the RNase P probe. Prime Script One-step First strand cDNA synthesis kit (Catalogue number: RR6110A, Takara Bio) along with gene specific primers (ORF1a and RNaseP) was used for cDNA preparation (as per manufacturers protocol). One step RT-PCR condition were 52° C for 5 minutes, 1 cycle (for reverse transcription), initial denaturation 95° C for 10 seconds followed by 40 cycles (for RT-PCR) of denaturation (95° C for 05 sec), (60° C for 90 sec) annealing/extension for PCR reaction.

#### 2.3 Assay Interpretation for Viroselect

The in-house Viroselect assay was considered positive if ORF1a (nsp3) and human RNase P (Internal Control) were detected. Negative, if ORF1a (nsp3) was not detected but RNase P was detected and invalid, if RNase P was not detected. Interpretation of STANDARD M nCoV Real-Time Detection kit (SD Biosensor, Gyeonggi-do, Republic of Korea) and LabGun (LabGenomics Co. Ltd., Gyeonggi-do, Republic of Korea) assays were as per manufacturers’ instructions.

#### 2.4 Interpretation for high throughput COBAS 6800 SARS-CoV-2 detection assay

As per the manufacturer, four types of interpretations were possible by the system (positive, negative, invalid, inconclusive). If target 1 (ORF), target 2 (E) and IC (non-infectious RNA in MS2 bacteriophage) were detected, the sample was interpreted as positive and if both targets were not detected but IC was detected, then the sample was interpreted as negative. If the samples showed target 1 negative, target 2 positive & IC positive, then the sample was interpreted as inconclusive.

### 3. Data analysis

Contingency Tables was constructed and positive and negative concordance along with over all agreement between various assays was determined.

## RESULTS

### Development of novel in-house Viroselect assay

Primers were designed using the SARS-CoV-2 reference genome available at NCBI genbank (Accession number: NC 045512). The primers and probe were uniquely designed for Viroselect assay from regions which had maximum variation with the SARS-CoV-2 and MERS-CoV-2. This hypervariable region was identified through NCBI align2 blast (Figure 1a). As shown in Table 1 the sequence targets for designing the primers and probe were present in ORF1a (nsp3) region of the SARS-CoV-2 genome between nucleotide positions 1933-3956. The internal control primer and probe were derived from human ribonuclease (RNase) P gene (GenBank accession number NM_ 006413) sequence (Emery et al., 2004). As shown in Figure 1a, the unique design of the primer can aid in differentiating between SARS-CoV and SARS-CoV-2 during mixed infection.

**Table 1:**
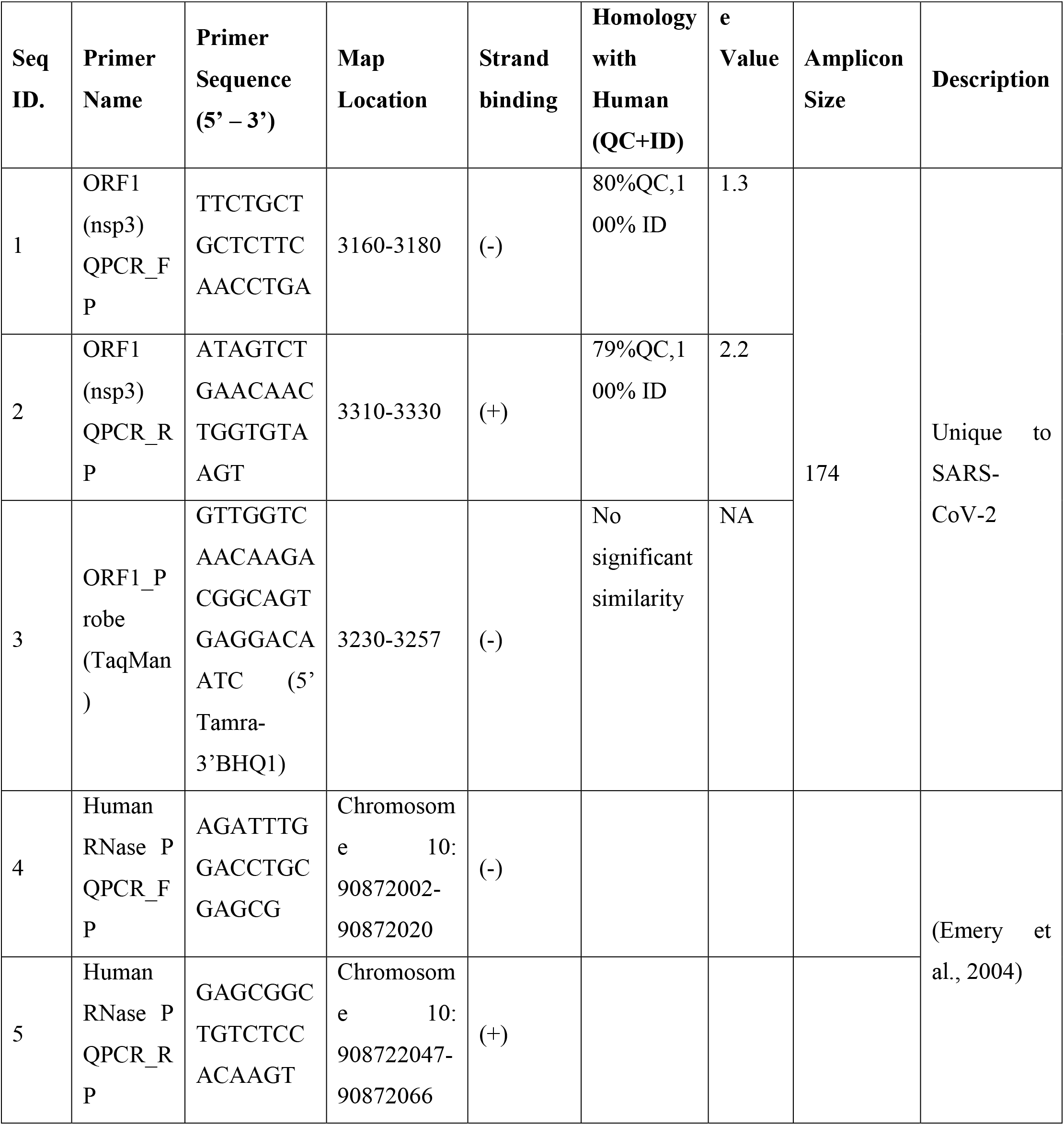
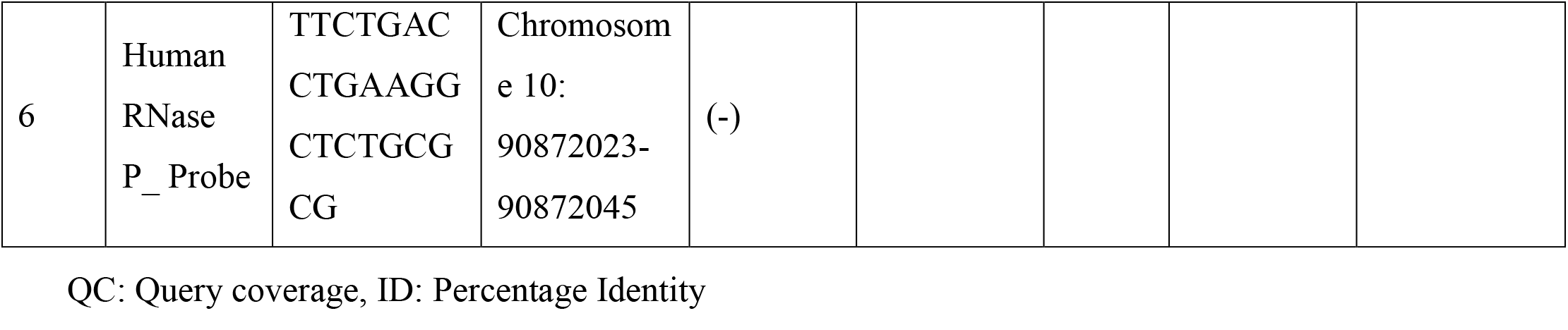
Primer sequences for SARS-CoV-2 detection (RT-PCR). Homology of ORF1a (*nsp3*) RT-PCR primers and Probe with Human Genome sprovided using nBLAST, highly similar sequences (megablast). QC: Query coverage, ID: Percentage Identity

**Figure 1a:**
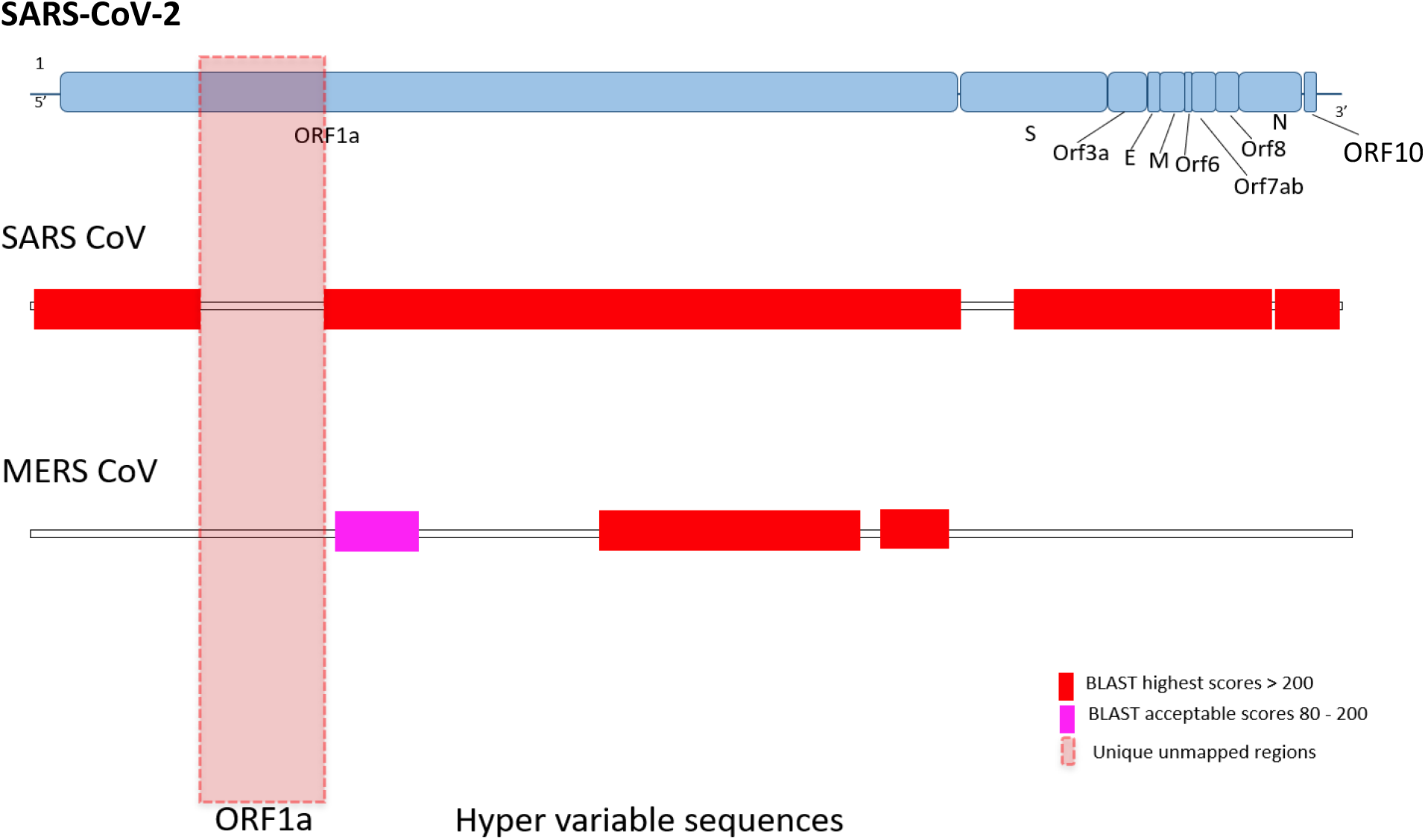
Depiction of the Hypervariable region between the genomes of SARS-CoV-2, SARS-CoV and MERS-CoV

NCBI nBlast was performed to analyze the sequence homology of putative amplicon with more than 250 SARS-CoV-2 isolates and 100% nucleotide sequence similarity was observed. Also, primers were designed to ensure that the putative amplicon did not overlap with known major mutation hotspots prevailing globally and in India (Figure 1b). Additionally, care was taken to have minimum homology with the human genome to avoid nonspecific amplification (Table 1). Following design of primers, amplicon was generated from infected samples and verified by agarose gel electrophoresis to be of the expected size (Figure 1c).

**Figure 1b:**
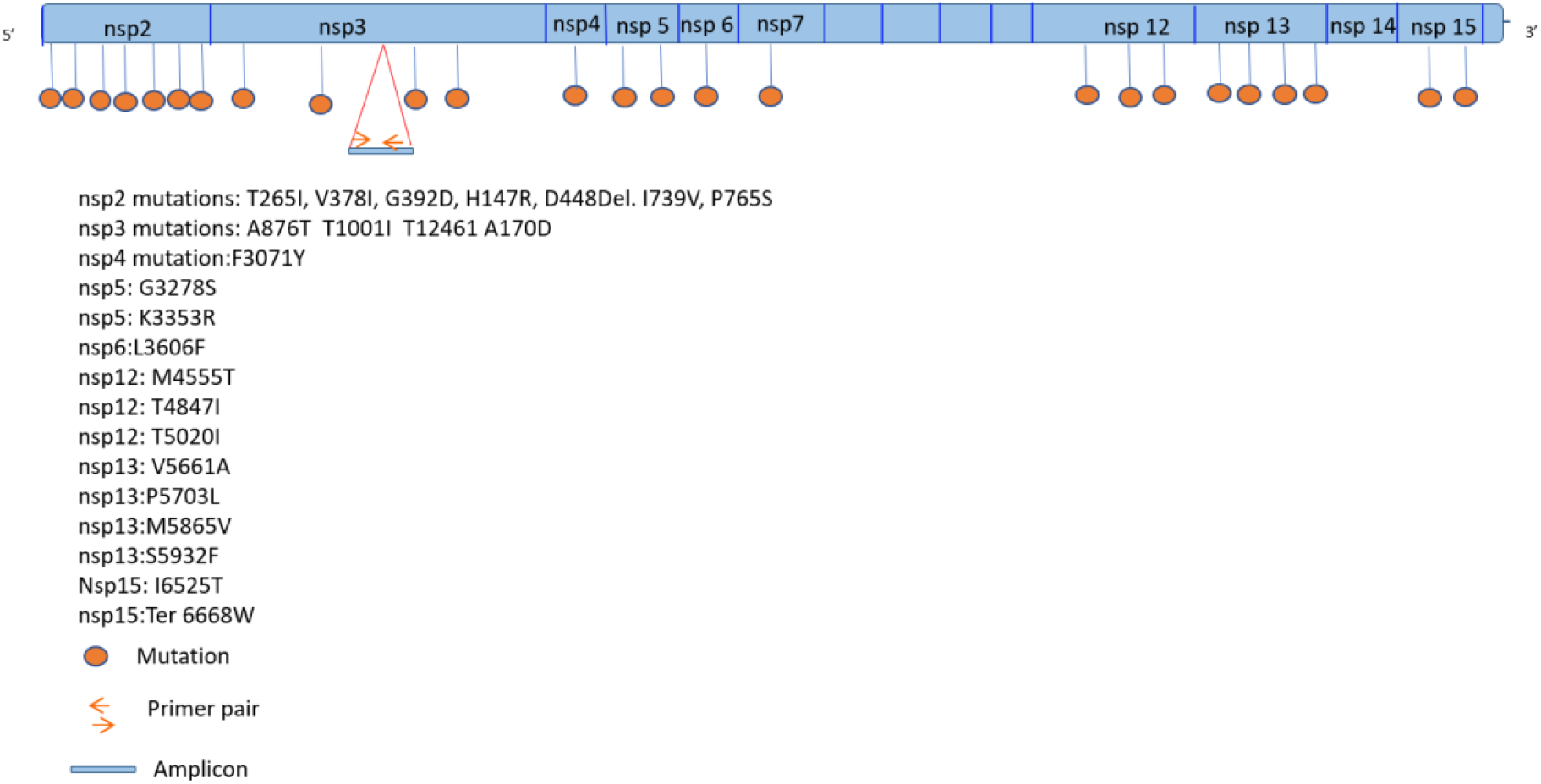
Depiction of primer position relative to major mutation hotspots of ORF1ab gene of SARS-CoV-2.

**Figure 1c:**
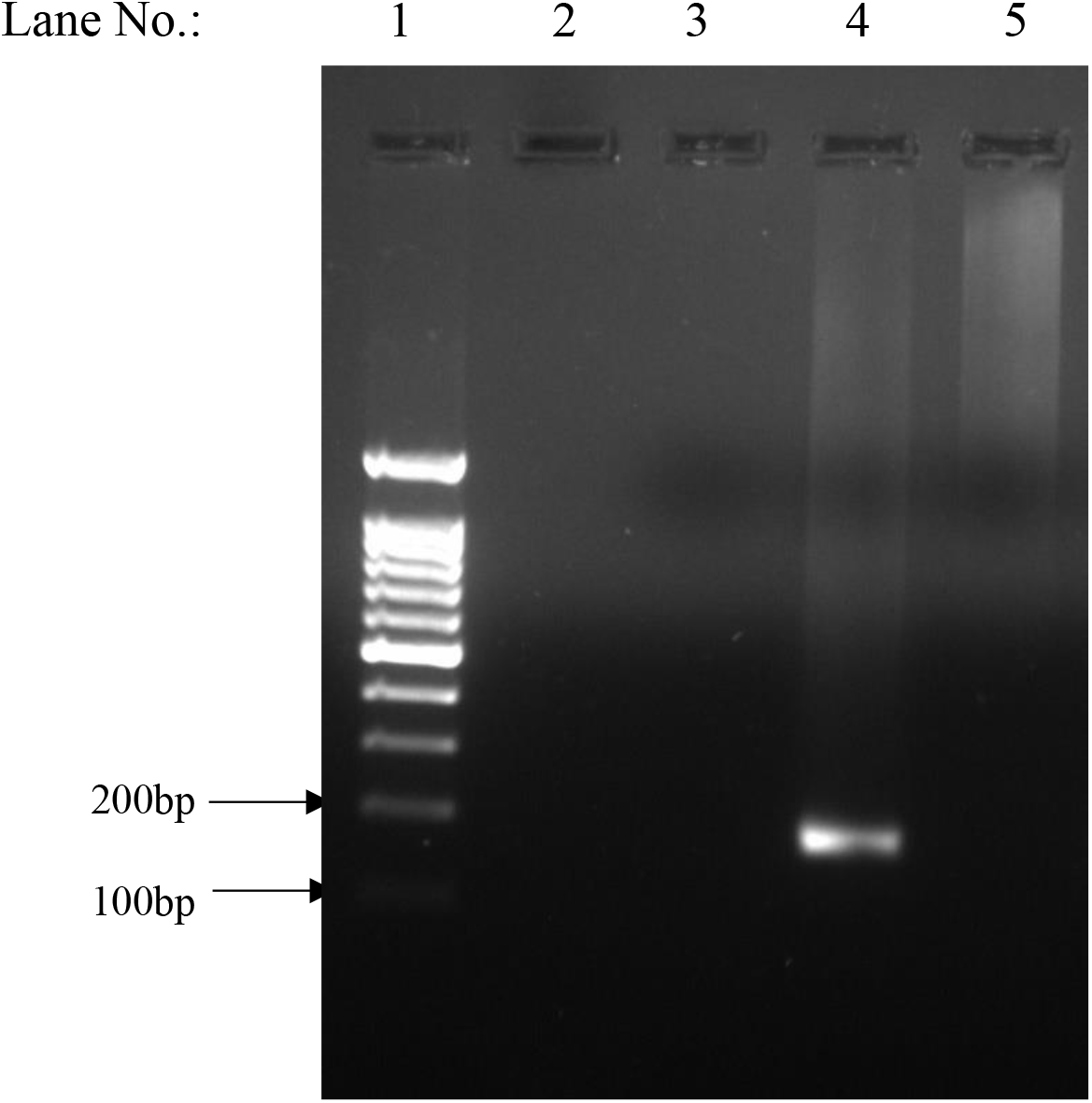
RT-PCR amplicon separated by 2% agarose gel electrophoresis after RNA extraction and 1^st^ strand cDNA synthesis. RNA extraction was carried out using QIAamp Viral RNA Mini Kit by QIAGEN and 1^st^ strand cDNA synthesis was carried out using Takara Prime Script 1^st^ strand cDNA synthesis kit. The PCR of the cDNA product was carried out using GeNei Taq DNA Polymerase kit and using the designed ORF1a forward and reverse primers. The expected amplicon size is 170bp which is observed in the figure. Lane 1: 100bp DNA Ladder (Marker) Lane 2: Blank Well Lane 3: Reagent Control Lane 4: COVID-19 Positive sample Lane 5: COVID-19 Negative sample

### Concordance Analysis

#### Viroselect with SD Biosensor assay

To assess the concordance of our in-house assay (Viroselect) with established and approved kits used for diagnosis, we evaluated the overall agreement of our assay with an EUA (USFDA) approved kit (SD Biosensor). RNA extraction from samples diagnosed as positive (n = 126) and negative (n = 107) for SARS-CoV-2 infection by SD Biosensor was followed by RT-PCR using Viroselect. Results revealed high concordance of the Viroselect assay with the SD Biosensor Kit as summarized in Table 2. As shown in Figure 2, a total of 286 samples (including 19 inconclusives) were tested of which 34 were invalid (not included in analysis). A concordance of 92.9% among positive samples and (81.3%) among negative samples was observed. An overall agreement of 87.6%, with 95% confidence interval (74.91% to 93.62%) was observed (Table 2). Interestingly a higher rate of discordance was observed in negative samples (20 out of 107; 18.7%) compared to that in positive samples (9 out of 126; 7.1%).

**Table 2:** Concordance of SD Biosensor positive and negative samples with Viroselect assay

|  | SD Biosensor |  |  | Concordance<br>among<br>positives | Concordance<br>among<br>negatives | Overall<br>agreement<br>(%) (95% CI) |
| --- | --- | --- | --- | --- | --- | --- |
| Viroselect<br>results | Positive | Negative | Total |  |  |  |
| Positive | 117 | 20 | 137 | 92.85% | 81.31% | 87.55%<br>(74.91 to<br>93.62) |
| Negative | 9 | 87 | 96 |  |  |  |
| Total | 126 | 107 | 233 |  |  |  |

**Figure 2:**
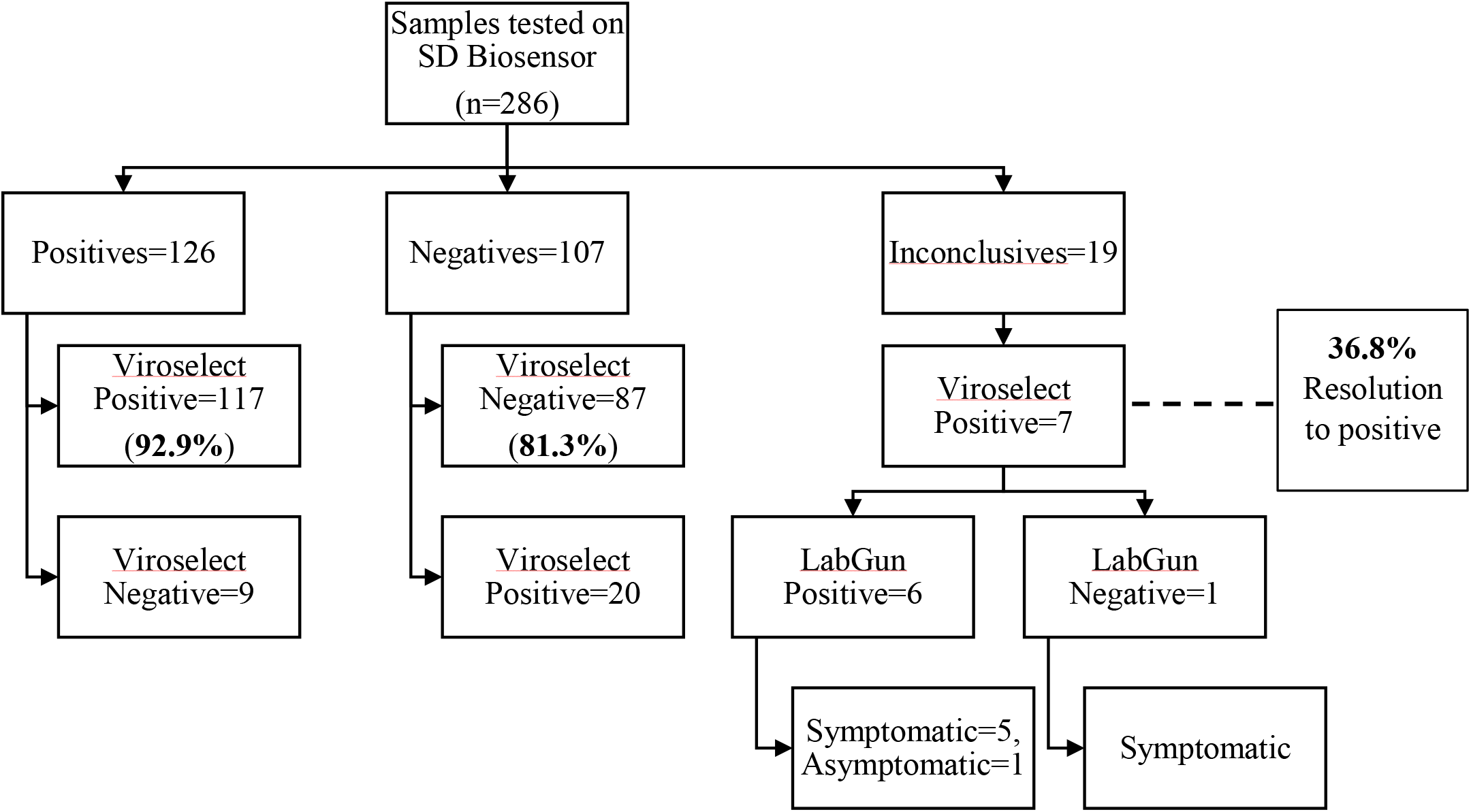
Flowchart showing Viroselect test results of the selected sample panel tested on SD Biosensor.

Notably, out of the 19 inconclusives by SD Biosensor kit, 7 (37%) were positive for Viroselect assay and 12 were negative. Of the 7 positives, six were positive when tested with another EUA (USFDA) approved RT-PCR kit, Labgun COVID-19 RT-PCR kit (LabGenomics). Further, clinical data obtained from the ICMR specimen referral form (SRF) revealed that six individuals out of seven that tested positive by Viroselect assay were symptomatic (Figure 2). Thus, Viroselect was able to resolve 100% of inconclusive samples tested by SD Biosensor.

#### Viroselect assay with COBAS 6800 High Throughput Platform

As our centre transitioned to a high throughput testing facility, we also had the opportunity to establish the concordance of Viroselect with SARS-CoV-2 detection assay on the COBAS 6800 High Throughput Platform (COBAS). Known COBAS positive (n= 102), negative (n=108) were tested by Viroselect. Seven of these samples were found to be invalid and not analysed further. Results revealed high concordance of the Viroselect assay with COBAS as summarized in Table 3. A concordance of 89% among positive samples and 84.5% among negative samples was observed with an overall agreement of 84.7%, with 95% confidence interval (76.00% to 90.85%) (Table 3). Interestingly, a higher rate of discordance was observed in negative samples (16 out of 103; 15.5%) compared to that in positive samples (11 out of 100; 11%).

**Table 3:**
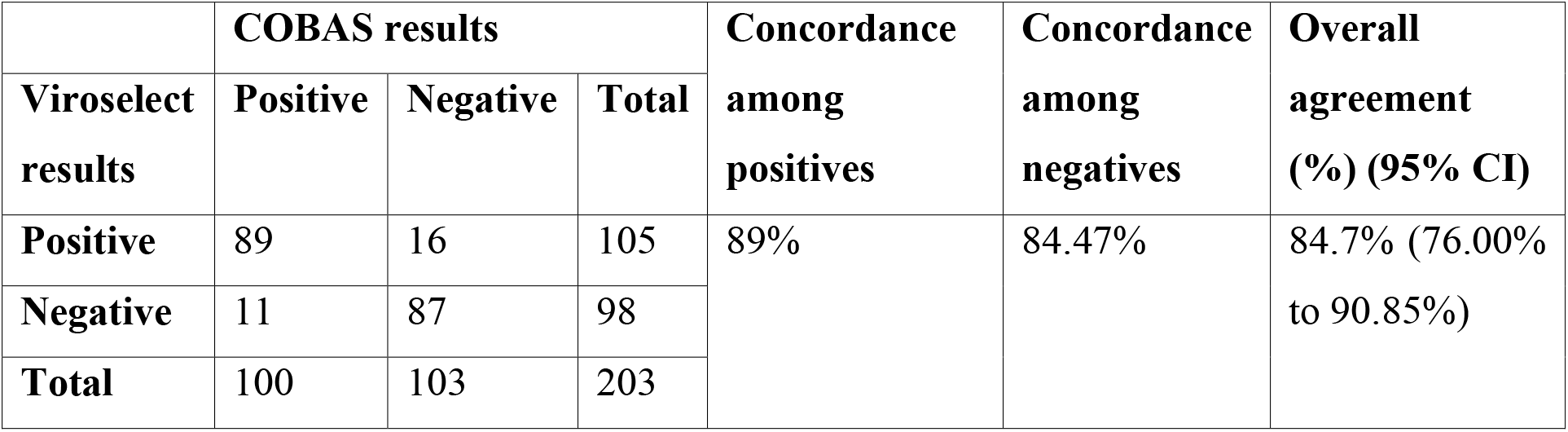
Concordance of COBAS positive and negative samples with Viroselect assay

|  | COBAS results |  |  | Concordance<br>among<br>positives | Concordance<br>among<br>negatives | Overall<br>agreement<br>(%) (95% CI) |
| --- | --- | --- | --- | --- | --- | --- |
| Viroselect<br>results | Positive | Negative | Total |  |  |  |
| Positive | 89 | 16 | 105 | 89% | 84.47% | 84.7% (76.00%<br>to 90.85%) |
| Negative | 11 | 87 | 98 |  |  |  |
| Total | 100 | 103 | 203 |  |  |  |

As shown in Figure 3, a total of 205 inconclusive samples were also tested of which 13 were invalid (not included in analysis). Of these, 175 (91.2%) tested negative and 17 (8.9%) tested positive. This indicates that 192 out of 205 (93.7%) of the COBAS inconclusive samples were resolved.

**Figure 3:**
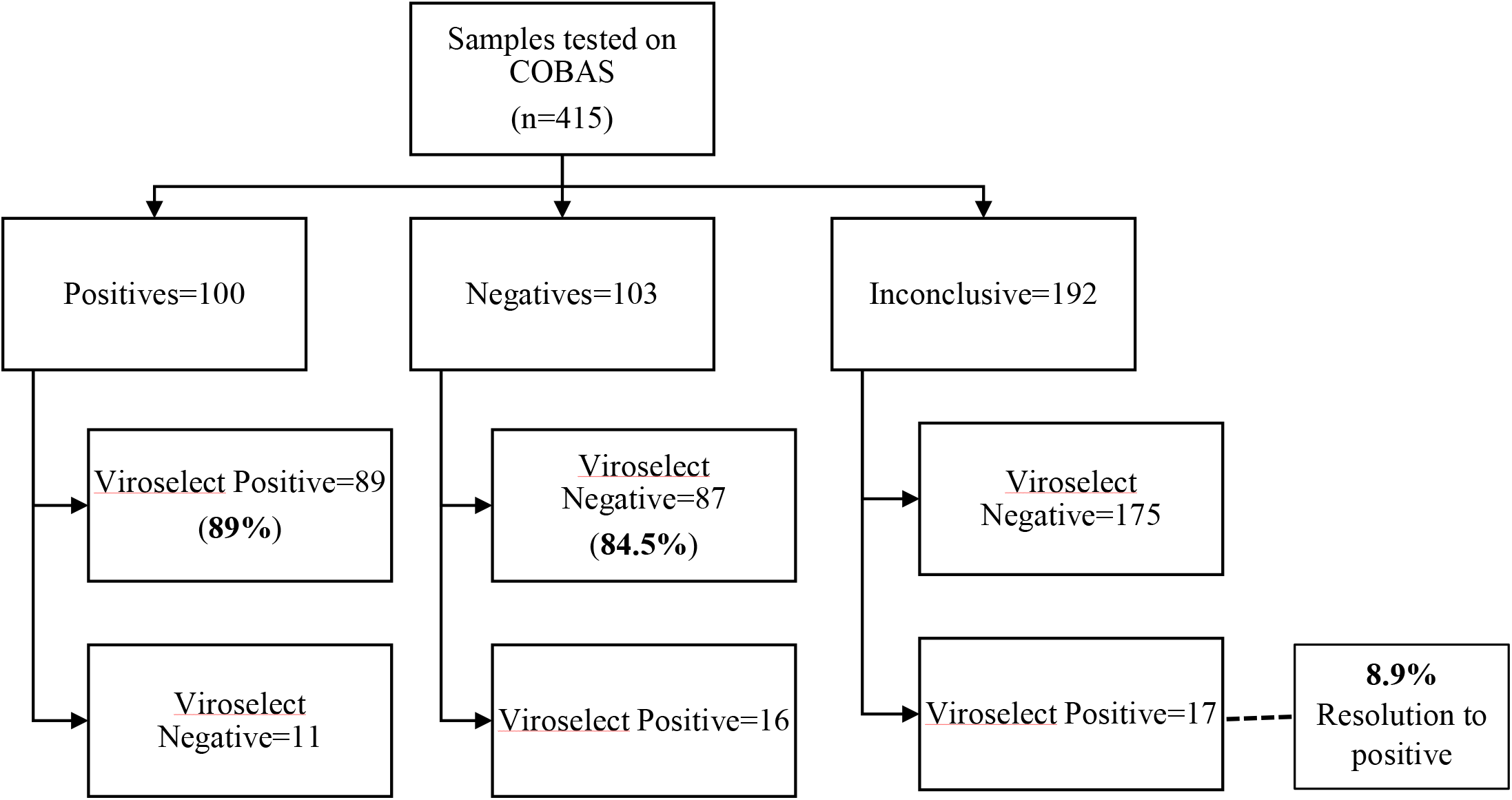
Flowchart showing Viroselect test results of the selected sample panel tested on COBAS 6800 platform.

## DISCUSSION

In this study, we demonstrate the utility of using a unique, ORF-1a based SARS-CoV-2 detection assay in resolving inconclusive results obtained from a conventional manual assay as well for those obtained from a high-throughput testing platform.

Real time PCR has been established as a gold standard for detecting SARS-CoV-2 infection. The whole genome sequencing of SARS-CoV-2 in a very large number, and availability of free databases (including, NCBI, GISAID) for accessing the genomes have attributed to development of multitude of RT PCR based kits which can often provide conflicting and inconclusive results (Fang et al., 2020). Thus, a testing strategy that would resolve these issues would greatly enhance efficiency of diagnosis which in tun would significantly impact transmission dynamics and allow efficient epidemiological surveillance. Also, the rapidly accumulating mutations in SARS-CoV-2 genome pose a challenge for accurately diagnosing the infection due to possible mispriming in assays that rely on detection of viral sequences in these regions (Rambaut et al., 2020). Based on hypervariable sequence analysis as well as mutation hotspot enumeration we designed unique primers capable of differentiating infection with SARS-CoV-2, SARS-CoV and MERS-CoV. The hypervariability was observed in the regions (1933-3956) of ORF1a and our primer/probe set spanned within the nsp3 region. This design strategy was implemented to develop an in-house RT-PCR based assay for detection of SARS-CoV-2.

Our unique primer and probe design were subjected to comparative analysis with 1) WHO recommended and EUA (USFDA) approved SD Biosensor kits, as well as with 2) the high throughput, automated SARS-CoV-2 detection assay on the COBAS 6800 platform. Overall high concordance was observed for our inhouse assay with both these detection modalities. Viroselect assay was able to resolve 100% of the inconclusive results obtained by the SD Biosensor kit. Notably, 7 out of 19 (36.8%) of these samples were resolved to be positive by our assay, while 6 out of 7 were resolved to be positive by another EUA (USFDA) approved kit, LabGun. Additionally, when corroborated clinically, 6 out of 7 inconclusive samples tested positive by Viroselect were documented to have been obtained from symptomatic individuals.

A precise and early diagnosis and treatment is crucial in COVID-19 (Rong et al., 2020). High-throughput diagnostic platforms, such as the COBAS 6800 are critical in addressing diagnostic demand during fast moving epidemics such as COVID-19 and have a direct impact on curtailing transmission as well as clinical management. Considering the similarity in geography, disease prevalence, population dynamics and healthcare spending of all the countries in South Asia, Ministry of Health and Family Welfare, Government of India adopted commendable measures to tackle the COVID-19 pandemic. A major such measure was increased accessibility to high throughput testing facilities. Our laboratory was one of these centres where we have tested close to 100,000 samples to date from Mumbai and surrounding areas. When dealing with such high throughput, a major challenge encountered was the rate of inconclusives. Although the rate was relatively low at 3%, the absolute number translated into approximately 3000 individuals (for every 100,000 individuals). Considering rapid transmission dynamics of SARS-CoV-2, even a day’s delay in correctly diagnosing and reporting a false negative or inconclusive sample as positive would result in significant impact on disease transmission in a highly populated and resource setting such as India (Kretzschmar et al., 2020; Larremore et al., 2021; Rong et al., 2020). When Viroselect was used to interrogate a subset of inconclusive samples obtained from our COBAS 6800 platform we found that our assay was able to successfully resolve ∼94% of these samples. Most importantly, ∼ 9% of these inconclusives were resolved to be positive. Our assay would thus enable optimal intervention in managing these cases, especially if they are assumed to be potential transmitters.

## Data Availability

All data is available on request

## Consent for publication

Not applicable

## Availability of data and materials

Not applicable

## Competing interests

The authors declare that they have no competing interests.

## Funding sources

This work was supported by Indian Council of Medical Research-National Institute for Research in Reproductive Health (ICMR-NIRRH). The funding body had no role in the design of the study, collection, analysis, and interpretation of data or in writing the manuscript.

## Authors’ contributions

DJ, SB1, and VP conceptualized the study. KJ, AYS, BC, SK, NK, SB2, SB3, PD, KK, HP performed the experiments. KJ, AYS, BC conducted primary data analysis and prepared the figures. JS, SA, KJ, BC and KM collected the clinical data. DJ, SB1, and VP supervised the study. VP and DJ wrote and edited the manuscript; all authors discussed the results and contributed to the final manuscript.

## Acknowledgments

We are grateful to Director General, Indian Council of Medical Research (ICMR), Ministry for Health & Family Welfare, Government of India for encouraging and supporting with resources, the pursuit of important research questions relevant to COVID-19 pathogenesis.

## Ethics approval

This study was approved by Institutional Ethics Committee at ICMR-NIRRH (Project No. 399-2020)

## Declaration of interests

⊠ The authors declare that they have no known competing financial interests or personal relationships that could have appeared to influence the work reported in this paper.

□The authors declare the following financial interests/personal relationships which may be considered as potential competing interestsxs:

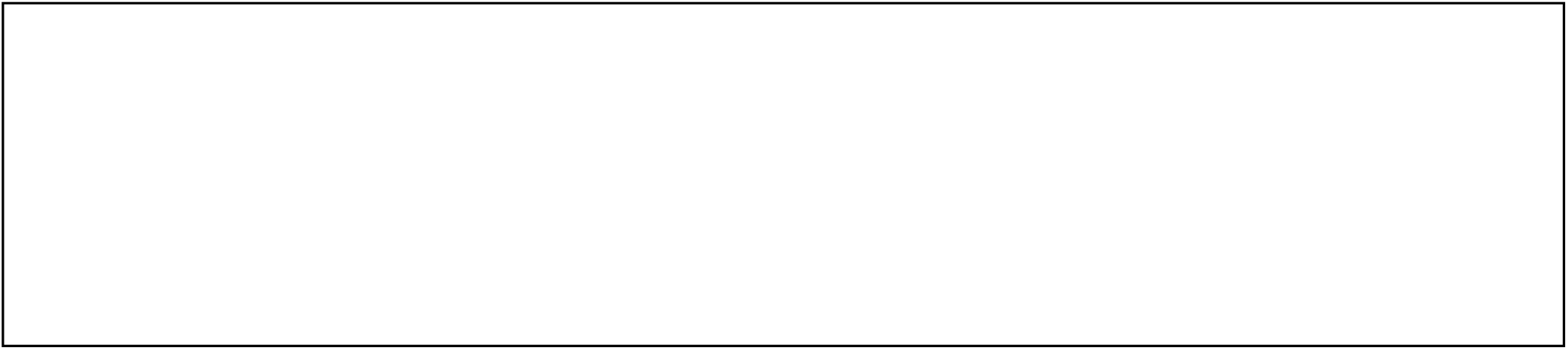

## Notes

### Competing Interest Statement

The authors have declared no competing interest.

### Funding Statement

ICMR-NIRRH Intramural Core Support

### Author Declarations

This study was approved by Institutional Ethics Committee at ICMR-NIRRH (Project No. 399-2020)

